# Early life modifiable risk factors for the development of knee osteoarthritis: protocol for a systematic review

**DOI:** 10.1101/2020.05.24.20112250

**Authors:** Ambrish Singh, Graeme Jones, Changhai Ding, Tania Winzenberg, Flavia Cicuttini, Swati Lavekar, Pablo Molina-Garcia, Petr Otahal, Benny Antony

## Abstract

Knee osteoarthritis (OA) is the most common form of OA which affects knee joints and there is currently no disease-modifying treatment available for OA. Therefore, an ideal strategy to prevent the development of OA is to identify and intervene at the modifiable risk factors for the development and progression of OA. Early-life factors such as obesity and malalignment may affect the mechanical aspect of the knee (i.e. alterations in normal knee kinematics) and could be the risk factor for the development of knee OA in later life. Identifying early-life (gestational factors, congenital defects, childhood, adolescence, early adulthood) factors which affect the development of knee OA in later stages of the life may help to develop targeted prevention programs in early-life itself to prevent the development of knee OA. Hence, this systematic review protocol provides the method to be used to comprehensively summarise the existing evidence on early life modifiable risk factors associated with the development and progression of knee OA.

## Review question

This systematic review aims to comprehensively summarise the existing evidence on early life modifiable risk factors associated with the development of knee OA.

## Searches

We will search MEDLINE® (via Ovid), EMBASE® (via Ovid), JBI Evidence-based Practice Database (via Ovid), Emcare (via Ovid), and Web of Sciences. Other resources to be searched include key conferences [(abstracts from last two years of European League Against Rheumatism (EULAR), Osteoarthritis Research Society International (OARSI), American Academy of Orthopaedic Surgeons (AAOS) and American College of Rheumatology (ACR))], bibliography of included studies, important systematic reviews and meta-analysis, and information about unpublished or incomplete trials from investigators known to be involved in previous important trials/studies.

## Types of study to be included

Observational studies such as. prospective, retrospective, cohort, case-control, and nested case-control studies will be included.

## Condition or domain being studied

Knee osteoarthritis (OA) is the most common form of OA which affects knee joints and there is currently no disease-modifying treatment available for OA.^1^ Therefore, an ideal strategy to prevent the development of OA is to identify and intervene at the modifiable risk factors for the development and progression of OA. Early-life factors such as obesity and malalignment may affect the mechanical aspect of the knee (i.e. alterations in normal knee kinematics) and could be the risk factor for the development of knee OA in later life.^2^ Identifying early-life (gestational factors, congenital defects, childhood, adolescence, early adulthood) factors which affect the development of knee OA in later stages of the life may help to develop targeted prevention programs in early-life itself to prevent the development of knee OA.^2^

## Participants/population

Participants (fetal stage, preterm neonatal, term neonatal, infancy, toddler, early childhood, middle childhood, early adolescence, late adolescence, early-adulthood) with a diagnosis confirmed as clinical knee OA, radiographic knee OA, or total knee replacement (TKR) due to knee OA.

## Intervention(s), exposure(s)

Any individual characteristic or event of early-life which is found to be associated with the increased likelihood of developing knee OA in later life will be considered as an early-life risk factor for knee OA.

## Comparator(s)/control

- A population who does not have the exposure (i.e. absence of risk factor).
- A population who doesn’t have the outcome (i.e. absence of knee OA/ TKR diagnosis).

## Primary outcomes

Incidence, prevalence, relative risk (RR), odds ratios (OR), or hazard ratio (HR) reported for the association between BMI/obesity measures and injury and development of knee OA and/or TKR due to knee OA will be extracted.

## Secondary outcomes

The secondary outcomes of interest will be (narrative synthesis planned):

1. Socioeconomic characteristics and risk of knee OA and/or TKR due to knee OA
2. Occupation/occupational characteristics and risk of knee OA and/or TKR due to knee OA
3. Joint pain, Joint diseases (Juvenile arthritis, Perthes’ disease, Osgood-Schlatter disease etc.), biomechanical markers and risk of knee OA and/or TKR due to knee OA
4. History of other diseases (e.g., not limited to, diabetes) and risk of knee OA and/or TKR due to knee OA
5. History of malalignment and risk of knee OA and/or TKR due to knee OA

## Data extraction (selection and coding)

Data will be extracted by two independent researchers using a standardised data extraction template: bibliometric data, study characteristics, characteristics of included participants, and outcome measures.

## Risk of bias (quality) assessment

The quality of the included studies will be assessed using the Joanna Briggs Institute Critical Appraisal Tool for Systematic Reviews.

## Strategy for data synthesis

We will summarize the study characteristics in the summary tables of the report and data will be presented as frequencies and percentages, means and standard deviations, or medians and interquartile ranges, as available. A meta-analysis of the outcome of interest will be performed using STATA version 16 (STATA Corp., Texas, USA) and Review Manager 5 (RevMan 5.3) (Copenhagen: The Nordic Cochrane Centre,

The Cochrane Collaboration, 2014). If a meta-analysis will not be possible considering heterogeneity in study designs, outcome variables, and reporting methods, a narrative synthesis will be developed to report the findings. In such a scenario, results will be presented in tables reporting individual risk factors (e.g., BMI, early life injury etc.).

## Analysis of subgroups or subsets

As per the availability of the data, we will perform the subgroup analysis or meta-regression analysis based on age group, gender and other subgroups.

## Data Availability

Not applicable

## APPENDIX 1. SEARCH STRATEGIES

MEDLINE® (via Ovid), EMBASE® (via Ovid), JBI Evidence-based Practice Database (via Ovid), Emcare (via Ovid)

| Review stage | Started | Completed |
| --- | --- | --- |
| Preliminary searches | No | No |
| Piloting of the study selection process | No | No |
| Formal screening of search results against eligibility criteria | No | No |
| Data extraction | No | No |
| Risk of bias (quality) assessment | No | No |
| Data analysis | No | No |

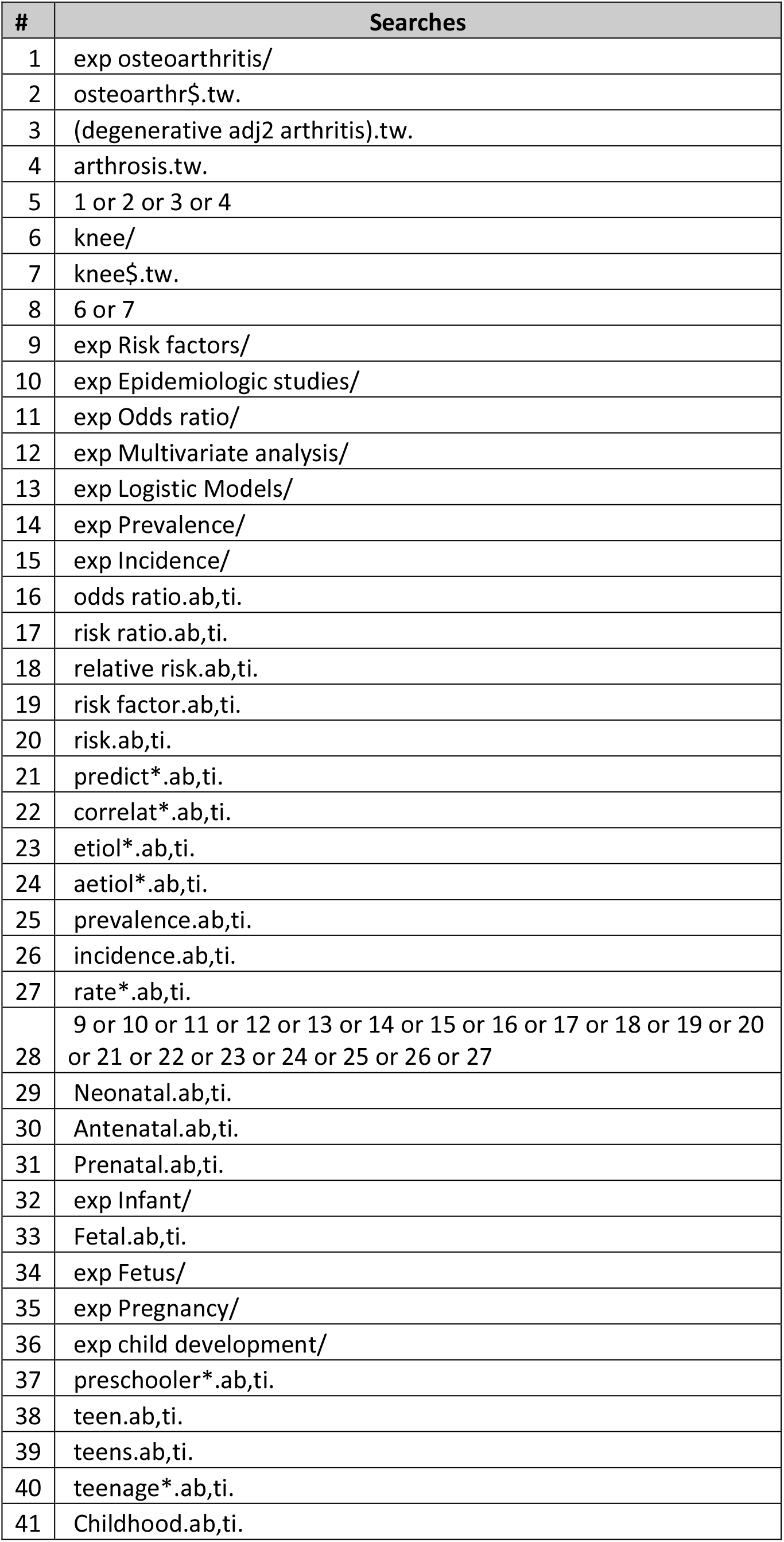

### Systematic review

#### 1. * Review title

Give the working title of the review, for example the one used for obtaining funding. Ideally the title should state succinctly the interventions or exposures being reviewed and the associated health or social problems. Where appropriate, the title should use the PI(E)COS structure to contain information on the Participants, Intervention (or Exposure) and Comparison groups, the Outcomes to be measured and Study designs to be included.

Early life modifiable risk factors for the development of knee osteoarthritis

#### 2. Original language title

For reviews in languages other than English, this field should be used to enter the title in the language of the review. This will be displayed together with the English language title.

#### 3. * Anticipated or actual start date

Give the date when the systematic review commenced, or is expected to commence.

20/05/2020

#### 4. * Anticipated completion date

Give the date by which the review is expected to be completed.

30/06/2021

#### 5. * Stage of review at time of this submission

Indicate the stage of progress of the review by ticking the relevant Started and Completed boxes. Additional information may be added in the free text box provided.

Please note: Reviews that have progressed beyond the point of completing data extraction at the time of initial registration are not eligible for inclusion in PROSPERO. Should evidence of incorrect status and/or completion date being supplied at the time of submission come to light, the content of the PROSPERO record will be removed leaving only the title and named contact details and a statement that inaccuracies in the stage of the review date had been identified.

This field should be updated when any amendments are made to a published record and on completion and publication of the review. If this field was pre-populated from the initial screening questions then you are not able to edit it until the record is published.

The review has not yet started: Yes

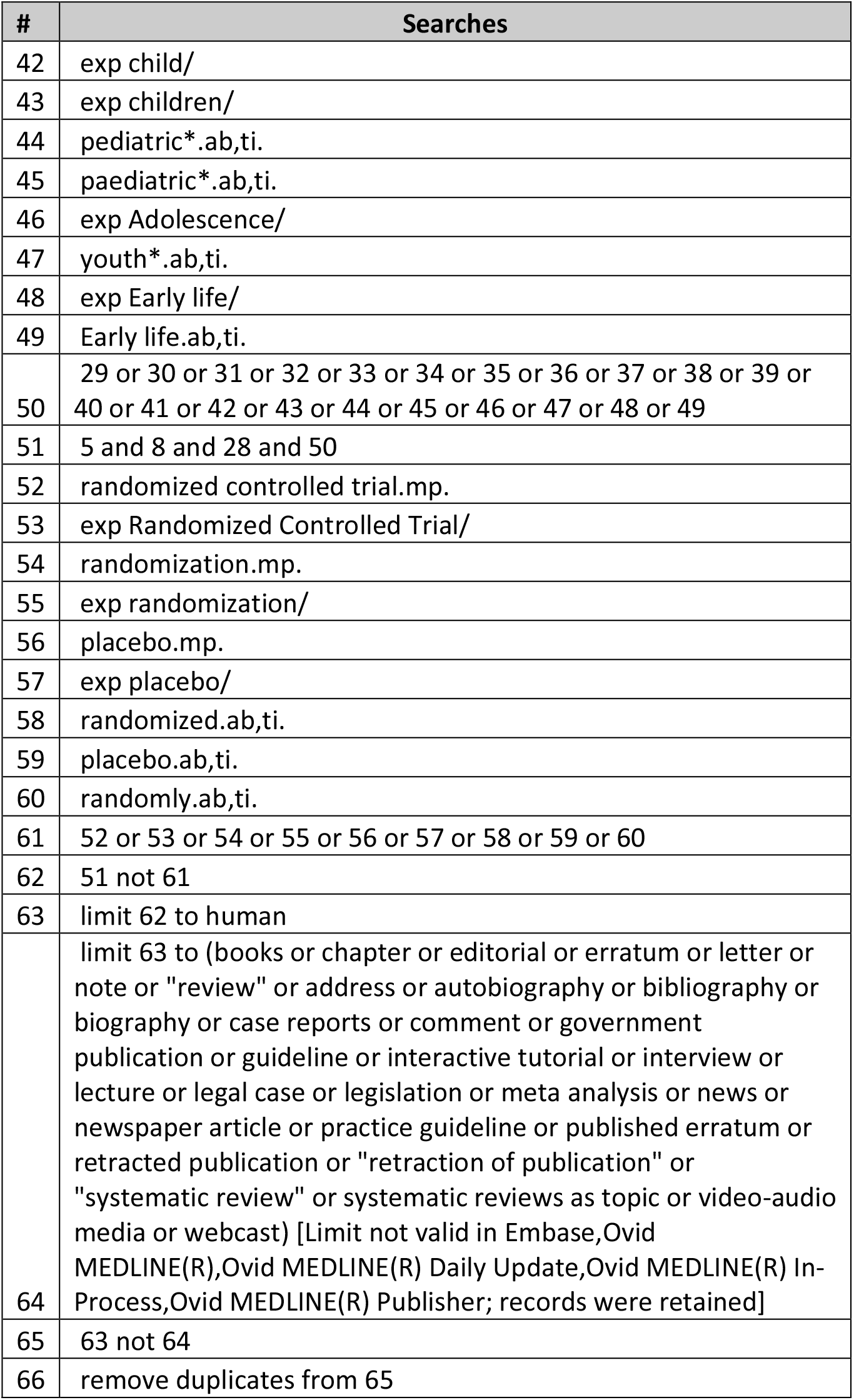

### PROSPERO

#### International prospective register of systematic reviews

Provide any other relevant information about the stage of the review here (e.g yet finalised).

This review will serve as a part of the PhD thesis.

This review will serve as a part of the PhD thesis.

#### 6. * Named contact

The named contact acts as the guarantor for the accuracy of the information presented in the register record. Ambrish Singh

Email salutation (e.g. “Dr Smith” or “Joanne”) for correspondence:

Mr Singh

#### 7. * Named contact email

Give the electronic mail address of the named contact.

#### 8. Named contact address

Give the full postal address for the named contact.

17 Liverpool St, Hobart TAS 7000

#### 9. Named contact phone number

Give the telephone number for the named contact, including international dialling code.

+610469701341

#### 10. * Organisational affiliation of the review

Full title of the organisational affiliations for this review and website address if available. This field may be completed as ‘None’ if the review is not affiliated to any organisation.

Menzies Institute for Medical Research, University of Tasmania, Hobart, Australia

#### Organisation web address: https://www.menzies.utas.edu.au/

#### 11. * Review team members and their organisational affiliations

Give the personal details and the organisational affiliations of each member of the review team. Affiliation refers to groups or organisations to which review team members belong. **NOTE: email and country are now mandatory fields for each person**.

Mr Ambrish Singh. Menzies Institute for Medical Research, University of Tasmania, Hobart, Australia Professor Graeme Jones. Menzies Institute for Medical Research, University of Tasmania, Hobart, Australia Professor Changhai Ding. Menzies Institute for Medical Research, University of Tasmania, Hobart, Australia Professor Tania Winzenberg. Menzies Institute for Medical Research, University of Tasmania, Hobart, Australia

Professor Flavia Cicuttini. Clinical Epidemiology, Monash University

Ms Swati Lavekar. Menzies Institute for Medical Research, University of Tasmania, Hobart, Australia Dr Pablo Molina-Garcia. PROFITH “PROmoting FITness and Health through physical activity” research group, Department of Physical Education and Sports, Faculty of Sport Sciences, University of Granada, Granada

Mr Petr Otahal. Menzies Institute for Medical Research, University of Tasmania, Hobart, Australia Dr Benny Antony. Menzies Institute for Medical Research, University of Tasmania, Hobart, Australia

#### 12. * Funding sources/sponsors

Give details of the individuals, organizations, groups or other legal entities who take responsibility for initiating, managing, sponsoring and/or financing the review. Include any unique identification numbers assigned to the review by the individuals or bodies listed.

This review received no funding other than salary stipends for authors as described below. BA is a recipient of the NHMRC Early-Career Fellowship, AS is a recipient of International Postgraduate Research Scholarship and GJ is a recipient of the NHMRC Practitioner Fellowship.

### Grant number(s)

#### 13. * Conflicts of interest

List any conditions that could lead to actual or perceived undue influence on judgements concerning the main topic investigated in the review.

None

#### 14. Collaborators

Give the name and affiliation of any individuals or organisations who are working on the review but who are not listed as review team members. **NOTE: email and country are now mandatory fields for each person**.

#### 15. * Review question

State the question(s) to be addressed by the review, clearly and precisely. Review questions may be specific or broad. It may be appropriate to break very broad questions down into a series of related more specific questions. Questions may be framed or refined using PI(E)COS where relevant.

#### 16. * Searches

State the sources that will be searched. Give the search dates, and any restrictions (e.g. language or publication period). Do NOT enter the full search strategy (it may be provided as a link or attachment.)

Relevant studies will be obtained from the following sources using the Ovid interface:

Ovid EMBASE®

Ovid JBI Evidence-based Practice Database Ovid Emc7are Web of Sciences

Searching other resources

- Conference search: We will search last two years abstracts from European League Against Rheumatism (EULAR), Osteoarthritis Research Society International (OARSI), American Academy of Orthopaedic Surgeons (AAOS) and American College of Rheumatology (ACR).
- We will hand search the bibliography of included articles and important systematic reviews and meta-analysis
- We will seek information about unpublished or incomplete trials from investigators known to be involved in previous important trials/studies.

#### 17. URL to search strategy

Give a link to a published pdf/word document detailing either the search strategy or an example of a search strategy for a specific database if available (including the keywords that will be used in the search strategies), or upload your search strategy.Do NOT provide links to your search results.

https://www.crd.york.ac.uk/PR0SPER0FILES/183272_STRATEGY_20200525.pdf

Alternatively, upload your search strategy to CRD in pdf format. Please note that by doing so you are consenting to the file being made publicly accessible.

Do not make this file publicly available until the review is complete

#### 18. * Condition or domain being studied

Give a short description of the disease, condition or healthcare domain being studied. This could include health and wellbeing outcomes.

Osteoarthritis (OA) is a chronic degenerative disease which mainly affects older populations; however, early life risk factors can have a role in the development of OA in later life. Knee OA is the most common form of OA which affects knee joints and there is currently no disease-modifying treatment available for knee OA. Moreover, early life factors such as obesity and malalignment may affect the mechanical aspect of the knee (i.e. alterations in normal knee kinematics) and could be the risk factor for the development of knee OA in later life. Identifying early life (gestational factors, congenital defects, childhood, adolescence, early adulthood) factors which affect the development of knee OA in later stages of the life may help to develop targeted prevention programs in early life itself to prevent the development of knee OA.

Hence, this systematic review aims to comprehensively summarise the existing evidence on early life modifiable risk factors associated with the development of knee OA.

#### 19. * Participants/population

Give summary criteria for the participants or populations being studied by the review. The preferred format includes details of both inclusion and exclusion criteria.

The population of interest will comprise participants, of age groups as described below, in whom the presence of one or more of the risk factor [such as BMI, obesity, occupational characteristics, physical activity, patellofemoral pain, injury, joint diseases (Juvenile arthritis, Perthes’ disease, Osgood-Schlatter disease etc.), biomechanical markers, other diseases (Diabetes etc.)] and its association on knee OA and/or total knee replacement (TKR) due to knee OA have been assessed.

Criteria to be used for the age group

Age stages according to NICHD pediatric terminology

Gestational: Fetal Stage

Preterm neonatal: the period at birth when a newborn is born before the full gestational period

Term neonatal: Birth– 27 d

Infancy: 28 d–2 mo

Toddler: 13 mo–2 y

Early childhood: 2-5 y

Middle childhood: 6-11 y Early adolescence: 12-18 y Late adolescence: 19-21 y Addition age group

Early-adulthood: Person between 21-30 years of age

#### 20. * Intervention(s), exposure(s)

Give full and clear descriptions or definitions of the nature of the interventions or the exposures to be reviewed.

Any modifiable individual characteristic or event of early life which is found to be associated with the increased likelihood of developing knee OA in later life will be considered as a modifiable early life risk factor for knee OA. Studies reporting data on modifiable risk factors associated with knee OA will be included. The modifiable risk factors may include, but not limited to, BMI, obesity, occupational, physical activity lifestyle, physical fitness level, patellofemoral pain, history of injury, joint diseases (juvenile arthritis, Perthes’ disease, Osgood-Schlatter disease etc.), biomechanical markers, and other diseases (Diabetes etc). These risk factors must be present at any stage of early life (as described in participants characteristic) and are present before the diagnosis of knee OA and/or TKR due to knee OA to confirm the temporality.

#### 21. * Comparator(s)/control

Where relevant, give details of the alternatives against which the main subject/topic of the review will be compared (e.g. another intervention or a non-exposed control group). The preferred format includes details of both inclusion and exclusion criteria.

For cohort studies: a population who does not have the exposure (i.e absence of risk factor).

For case-control studies: a population who doesn’t have the outcome (i.e absence of knee OA and/or TKR due to knee OA diagnosis).

#### 22. * Types of study to be included

Give details of the types of study (study designs) eligible for inclusion in the review. If there are no restrictions on the types of study design eligible for inclusion, or certain study types are excluded, this should be stated. The preferred format includes details of both inclusion and exclusion criteria.

We will include observational studies (prospective, retrospective, cohort, case-control, and nested case-control studies).

We anticipate a very large number of articles to be included, hence we will exclude from this review case reports/case studies (less than 30 cases). The editorials, literature reviews, viewpoints/opinion, recommendations, guidelines, qualitative research, consensus papers, and grey literature, such as reports, thesis, and notes will also be excluded.

#### 23. Context

Give summary details of the setting and other relevant characteristics which help define the inclusion or exclusion criteria.

Studies from any setting assessing the association between preterm neonatal, childhood, adolescence, early adulthood modifiable factors and the development of knee OA and/or TKR due to knee OA will be included.

#### 24. * Main outcome(s)

Give the pre-specified main (most important) outcomes of the review, including details of how the outcome is defined and measured and when these measurement are made, if these are part of the review inclusion criteria.

The outcomes will be defined based on clinical knee OA, radiographic knee OA, or total knee replacement (TKR) due to knee OA.

We aim to identify the early life factors which are associated with knee OA and/or TKR due to knee OA.

The primary outcomes of interest will be (meta-analysis planned):

1. Early life BMI/obesity and risk of knee OA and/or TKR due to knee OA
2. Early life injury and risk knee OA and/or TKR due to knee OA

#### * Measures of effect

Please specify the effect measure(s) for you main outcome(s) e.g. relative risks, odds ratios, risk difference, and/or ‘number needed to treat.

Incidence, prevalence, relative risk (RR), odds ratios (OR), or hazard ratio (HR) reported for the association between obesity measures and development of knee OA and/or TKR due to knee OA will be extracted.

#### 25. * Additional outcome(s)

List the pre-specified additional outcomes of the review, with a similar level of detail to that required for main outcomes. Where there are no additional outcomes please state ‘None’ or ‘Not applicable’ as appropriate to the review

The secondary outcomes of interest will be (narrative synthesis planned):

1. Socioeconomic characteristics and risk of knee OA and/or TKR due to knee OA
2. Occupation/occupational characteristics and risk of knee OA and/or TKR due to knee OA and/or TKR due to knee OA
3. Joint pain, Joint diseases (Juvenile arthritis, Perthes’ disease, Osgood-Schlatter disease etc.), biomechanical markers and risk of knee OA and/or TKR due to knee OA
4. History of other diseases (e.g., not limited to, diabetes) and risk of knee OA and/or TKR due to knee OA
5. History of malalignment and risk of knee OA and/or TKR due to knee OA

#### * Measures of effect

Please specify the effect measure(s) for you additional outcome(s) e.g. relative risks, odds ratios, risk difference, and/or ‘number needed to treat.

Incidence, prevalence, relative risk (RR), odds ratios (OR), or hazard ratio (HR) reported for the development of knee OA and/or TKR due to knee OA will be extracted.

#### 26. * Data extraction (selection and coding)

Describe how studies will be selected for inclusion. State what data will be extracted or obtained. State how this will be done and recorded.

Selection of studies

Two review authors will independently screen the titles and abstracts of search output and classify them as ‘eligible/potentially eligible for inclusion’ or ‘not eligible’. All the studies classified as eligible based on title and abstract will be retrieved and screened based on full-text by two review authors independently. Any disagreement will be resolved by discussion or consultation with senior authors.

Data extraction and management

Data extraction will be performed by two independent reviewers using an MS excel based data extraction template. We will extract the following data from each eligible studies: study author, study design, setting, follow-up duration, mean age of the participants, gender, ascertainment of exposure (risk factor) and outcomes (diagnosis of knee OA and/or TKR due to knee OA) etc. Any disagreement in data extraction from the included studies will be resolved by discussion or by involving the senior authors.

Risk factors will be classified into patient variables, disease characteristics, and biochemical or imaging markers. The outcomes will be classified into clinical knee OA, radiographic knee OA, or TKR due to knee OA.

#### 27. * Risk of bias (quality) assessment

Describe the method of assessing risk of bias or quality assessment. State which characteristics of the studies will be assessed and any formal risk of bias tools that will be used.

The quality of the included studies will be assessed using the Joanna Briggs Institute Critical Appraisal Tool for Systematic Reviews. The tool consists of several items which will be marked as: V (criterion met), ‘X’ (criterion not met) and ‘NA’ (not applicable). We will calculate the quality score for each study and considered as ‘high quality’ when the quality score is at least 0.75 (i.e., 75%), whereas studies would be considered as ‘low quality’ when the quality score is lower than 0.75. Quality assessment will be performed independently by two reviewers and disagreements would be discussed to reach the consensus.

#### 28. * Strategy for data synthesis

Provide details of the planned synthesis including a rationale for the methods selected. This **must not be generic text** but should be **specific to your review** and describe how the proposed analysis will be applied to your data.

We will summarize the study characteristics in the summary tables of the report and data will be presented as frequencies and percentages, means and standard deviations, or medians and interquartile ranges, as available.

We will assess the feasibility of meta-analysis based on the similarity of risk factors, clinical population, type of outcomes, and study designs. If appropriate we will use a random-effects model to meta-analyze the study outcomes, as we anticipate the included studies to be heterogeneous. For dichotomous outcomes, we will compute/extract the risk ratios, odds ratios, hazard ratios, with 95% confidence intervals (CIs), as available. For the continuous outcomes, mean or standardized mean differences with 95% CIs will be computed/extracted, as available. We will assess the statistical heterogeneity using the I^2^ statistics with I^2^ value of (0–25%), (25–50%), (50–75%), and (75–100%), denoting low, moderate, substantial, and considerable heterogeneity, respectively. We will use funnel plots to assess for publication bias if more than 10 studies are combined for meta-analysis. We will perform the statistical analysis using STATA version 16 (STATA Corp., Texas, USA) and Review Manager 5 (RevMan 5.3) (Copenhagen: The Nordic Cochrane Centre, The Cochrane Collaboration, 2014).

If a meta-analysis will not be possible considering heterogeneity in study designs, outcome variables, and reporting methods, a narrative synthesis will be developed to report the findings. In such a scenario, results will be presented in tables reporting individual risk factors (e.g., BMI, early life injury etc.).

#### 29. * Analysis of subgroups or subsets

State any planned investigation of ‘subgroups’. Be clear and specific about which type of study or participant will be included in each group or covariate investigated. State the planned analytic approach.

Based on the availability of the data, we will perform the subgroup analysis or meta-regression analysis, as appropriate, based on age group, gender and other subgroups.

#### 30. * Type and method of review

Select the type of review and the review method from the lists below. Select the health area(s) of interest for your review.

#### Type of review

Cost effectiveness No

Diagnostic

No

Epidemiologic

Yes

Individual patient data (IPD) meta-analysis

No

Intervention

No

Meta-analysis

Yes

Methodology

No

Narrative synthesis

No

Network meta-analysis

No

Pre-clinical

No

Prevention

No

Prognostic

Yes

Prospective meta-analysis (PMA)

No

Review of reviews

No

Service delivery

No

Synthesis of qualitative studies

No

Systematic review

Yes

Other

No

#### Health area of the review

Alcohol/substance misuse/abuse

No

No

Cancer

No

Cardiovascular

No

Care of the elderly

No

Child health

No

Complementary therapies

No

COVID-19

No

Crime and justice

No

Dental

No

Digestive system

No

Ear, nose and throat

No

Education

No

Endocrine and metabolic disorders

No

Eye disorders

No

General interest

No

Genetics

No

Health inequalities/health equity

No

Infections and infestations

No

International development

No

Mental health and behavioural conditions

No

Musculoskeletal

Yes

Neurological

No

Nursing

No

Obstetrics and gynaecology

No

Oral health

No

Palliative care

No

Perioperative care

No

Physiotherapy

No

Pregnancy and childbirth

No

Public health (including social determinants of health)

No

Rehabilitation

No

Respiratory disorders

No

Service delivery

No

Skin disorders

No

Social care

No

Surgery

No

Tropical Medicine

No

Urological

No

Wounds, injuries and accidents

No

Violence and abuse

No

#### 31. Language

Select each language individually to add it to the list below, use the bin icon to remove any added in error. English

There is not an English language summary

#### 32. * Country

Select the country in which the review is being carried out from the drop down list. For multi-national collaborations select all the countries involved.

Australia

#### 33. Other registration details

Give the name of any organisation where the systematic review title or protocol is registered (such as with The Campbell Collaboration, or The Joanna Briggs Institute) together with any unique identification number assigned. (N.B. Registration details for Cochrane protocols will be automatically entered). If extracted data will be stored and made available through a repository such as the Systematic Review Data Repository (SRDR), details and a link should be included here. If none, leave blank.

#### 34. Reference and/or URL for published protocol

Give the citation and link for the published protocol, if there is one Give the link to the published protocol.

Alternatively, upload your published protocol to CRD in pdf format. Please note that by doing so you are consenting to the file being made publicly accessible.

No I do not make this file publicly available until the review is complete

Please note that the information required in the PROSPERO registration form must be completed in full even if access to a protocol is given.

#### 35. Dissemination plans

Give brief details of plans for communicating essential messages from the review to the appropriate audiences.

This review will serve as a part of the PhD thesis of AS, who is enrolled as a PhD student under the senior investigators in this review. This review will be submitted to peer review journals for publication.

### Do you intend to publish the review on completion?

Yes

#### 36. Keywords

Give words or phrases that best describe the review. Separate keywords with a semicolon or new line. Keywords will help users find the review in the Register (the words do not appear in the public record but are included in searches). Be as specific and precise as possible. Avoid acronyms and abbreviations unless these are in wide use.

Childhood; BMI; Overweight; Exercise; Fitness; Physical activity; Adulthood; Knee pain; Osteoarthritis

#### 37. Details of any existing review of the same topic by the same authors

Give details of earlier versions of the systematic review if an update of an existing review is being registered, including full bibliographic reference if possible.

This systematic review will be an update to the narrative review done by senior author of this review. Citation for existing review, Antony B, et al. Do early life factors affect the development of knee osteoarthritis in later life: a narrative review. Arthritis Res Ther. 2016;18(1):202.

#### 38. * Current review status

Review status should be updated when the review is completed and when it is published. For newregistrations the review must be Ongoing.

Please provide anticipated publication date

Review_Ongoing

#### 39. Any additional information

Provide any other information the review team feel is relevant to the registration of the review.

#### 40. Details of final report/publication(s) or preprints if available

This field should be left empty until details of the completed review are available OR you have a link to a preprint.

Give the link to the published review.

